# Breast Cancer Detection Using Quantum Convolutional Neural Networks: A Demonstration on a Quantum Computer

**DOI:** 10.1101/2020.06.21.20136655

**Authors:** Aradh Bisarya, Walid El Maouaki, Sabyasachi Mukhopadhyay, Nilima Mishra, Shubham Kumar, Bikash K. Behera, Prasanta K. Panigrahi, Debashis De

## Abstract

Deep learning have paved the way for scientists to achieve great technical feats. In an endeavor to hone and perfect these techniques, quantum deep learning is a promising and important tool to utilize to the fullest. Using the techniques of deep learning and supervised learning in a quantum framework, we are able to propose a quantum convolutional neural network and showcase its implementation. Using the techniques of deep learning and supervised learning in a quantum framework, we are able to propose a quantum convolutional neural network and showcase its implementation. We keep our focus on training of the ten qubits system in a way so that it can learn from labeling of the breast cell data of Wisconsin breast cancer database and optimize the circuit parameters to obtain the minimum error. Through our study, we also have showcased that quantum convolutional neural network can outperform its classical counterpart not only in terms of accuracy but also in the aspect of better time complexity.

## I. INTRODUCTION

Techniques to mimic human intelligence using logic, algorithms, and networks are state of the art findings of science and technology. Artificial intelligence, along with its subsets of machine learning and deep learning, takes a giant leap in analyzing and classifying data. Neural networks prove to be the perfect programming-paradigm in this field [1–3]. With deep learning as its backbone, these deep neural networks are proficient in areas such as speech recognition, image recognition, data analysis, and much more.

Deep learning is based on the idea of managing the input data set through several layers of representation [4]. A layer can be described by a set of parameters, which take an input set and generate a decision or output. Deep learning is used to determine the appropriate parameters for a given application. The learning can be supervised or unsupervised. In supervised learning, which we have employed, we try to set the parameters from the input training set. In this learning method, each input training set is labeled with the desired output value. Thus, during the training phase, the system has been told what the output should be for the given input. In unsupervised learning, the data is analyzed without its labels so the program does not know what the output should look like, and grows slowly and more aimlessly. Unsupervised learning is mostly used to understand the data better. This entire process of deep learning is inspired by the functioning of the human brain and referred as the complicated form of neural networks.

Amalgamating convolutional neural network (CNN) models with concepts of quantum information and quantum mechanics, we are able to propose a quantum convolutional neural network (QCNN) model. Training the deep neural networks with data analysis features means that quantum computing based deep neural networks get an edge over artificial neural networks. However, quantum convolutional neural network models are mostly theoretical proposals as their full implementation in the physical world is yet to be explored [3].

In this paper, we attempt to build a quantum convolutional neural network model, exhibiting the use of several algorithms to handle the data set. Out of many applications of deep neural networks, we have taken breast cancer detection into account [5, 6]. An attempt to train the network to calculate the label function and minimize the loss function using standard algorithms has been made. We have tried to detect breast cancer on the basis of the label found from the input data set. Initially, we tried to give an insight into the process of detection of the disease by the image recognition method. Later, to gain the greater success, we implement the deep learning algorithm in quantum framework like quantum convolutional neural network (QCNN) along with its classical counterpart i.e, CNN on our input data set consisting of parameters associated with the features like cell sizes and textures. The classification results obtained are quite convincing demonstrating the application of the quantum convolutional neural network (QCNN) in comparisons to classical CNN algorithm. We further utilized the effectiveness of quantum CNN over classical CNN in terms of optimum time complexity while predicting the breast cancer cells. This work is a step taken to showcase the efficacy of a quantum deep neural network like QCNN.

IBM quantum experience, due to its easy and free access to the cloud-based quantum computer, has become a large platform for the research community in the field of quantum computation and quantum information to accomplish various tasks such as quantum simulation [7–9], quantum algorithms [10], quantum information-theoretical demonstrations [11, 12], quantum machine learning [13, 14], quantum error correction [15], quantum applications [16–18] to name a few. Here, we use this platform for the task of implementing QCNN as an application of breast cancer detection.

## II. METHODS

Deep learning along with supervised and unsupervised learning have given an edge over the other methods while handling the data. The product of these techniques is the architecture of a neural network. These artificial neural networks, through their layered structure, help the system to learn data handling. The input set of data, usually known as the training data, comes with some label value. It is then processed through the layers of the network such that the output label obtained is close to the true label. This basic framework is then extended to applications of image and pattern recognition, text classification, speech recognition and many more.

A quantum convolutional neural network (QCNN), capable of operating on a 10-qubit system, is designed. The designed QCNN is capable of carrying out several algorithms for optimizations. This design helps in obtaining the training vector to calculate the label value of the string given as input. Demonstrating the implementation of the network, we have tried to use the basic principles of deep learning to handle data, for detecting breast cancer. We have analysed two methods here. First by training the network through the numerical dataset and secondly, tried to utilize the method of image recognition. All the network implementation and simulation of results were carried out on the cloud-based platform provided by IBM.

### A. Process of Image Recognition

Demonstrating the idea of using image recognition as a tool to detect the disease on a quantum convolutional neural network.

#### Background

The process of detection of a disease such as breast cancer can be carried out through the method of image recognition. The image is analysed as a gray scale 2-dimensional matrix, whose cell value corresponds to relative brightness.

#### The quantum setting of the process

The process of image recognition is based on the grey scale data of the image. The system assigns values to each pixel, ranging from 0 to 1 based on its brightness. These values are then fed as the input training set in the form of an array. We also input the true label (whether the cell is cancerous or not) for the image in the training stage, which is either 0 or 1. The network is then trained to find the label value of the input set. This will be a value between 0 and 1, and the error is calculated based on its deviation from the true label. We collect the data set, i.e., images from Kaggle, which consist of the MRI images of the cells. The process involves entering the pixel matrix as the input to the neural network.

In theory, we may consider such a system to be built. However,for a reasonably high resolution image, the size of the matrix is too large to be given as input to the currently available quantum simulator circuit. As a test of a modified system, we chose a 4 *×* 4 matrix of pixels from the image and enter it as the dataset. The training parameter is thus calculated which gives the minimum error or loss. To minimize the loss function, we have implemented the optimization algorithm like variational quantum eigensolver (VQE). We are able to determine the training parameter which gives the minimum error while calculating the label function. The parameters are thus chosen which are optimal to the given problem.

The matrix that is sent as an input is selected from the image. A validation set is obtained which is an array of size 4 *×* 4, taken randomly from the image. This is then coded into the network for classification and to obtain the label value of the training set. However, as mentioned, an entire 256 qubits system could not be simulated owing to the unavailability of the required system. We act on the input set using rotation gates of Z gates and X gates to gradually change the training parameter following optimization to give the minimum deviation of the obtained label value from the true label value. The simulation is carried using the API based cloud access provided by IBM. The method, however, gives down-sampled results owing to the random selection of apart from the image and not sending the entire picture, due to lack of required system. Though this algorithm can be more aptly implemented upon the availability of a higher qubit handling a quantum computer.

### B. Using the data set consisting of information on cell features

The detection of breast cancer in early stages has been done by studying subtle morphological changes due to broken collagen fibers for disease progression. The MRI cell dataset of Wisconsin Breast Cancer consists of features of lumps on the patient’s breast, such as, radius, roughness, concavity, and so on. The data was taken from the cited source [19]. We detect whether the person suffers from breast cancer using the data providing information on the size, radius, etc. of the affected cell, by implementing a QCNN. This method has the advantage of having a smaller initial data set and being implemented on currently available simulators.

#### Background

Classification of data has been an important part of machine learning. The principles of data classification and deep learning can aid us to detect breast cancer in a woman patient. We, thus, design a quantum convolutional neural network and demonstrate its implementation by detecting breast cancer in the patient.

#### The quantum setting of the process

Analogous to neurons and weights in a classical deep learning neural network, we have qubits and their connections along with the rotation gates. The design is carried on a 10-qubits system. Our main intention is to obtain the training parameter that implies to minimum deviation of the obtained label value from its true value. Each string as an input has some label value assigned to it. The inputs are fed to the circuit having some rotation parameters. Following the approach in deep learning algorithms, the training vector is varied after every iteration to correspond to minimum loss. The optimization algorithm of online gradient descent has been implemented. The rate of decrease is also regulated. This method gives successful interpretations on the operation of the network giving further results on the analysis of variation of training parameters and loss functions. The circuit design and implementation is carried through the IBM quantum experience.

## III. ARCHITECTURE OF QCNN

Neural networks may be implemented in quantum computers by treating the qubits as neurons. We act on neurons with rotation gates which have parameters, which are then optimized in our model. The inputs are implemented as initial rotations on each qubit. The operational block diagrams of classical CNN and quantum CNN on breast cell samples are shown in Fig 1(a) and 1(b). Let us make a comparison based on the equivalence between CNN and QCNN to understand the structure of the QCNN circuit:

**FIG. 1:**
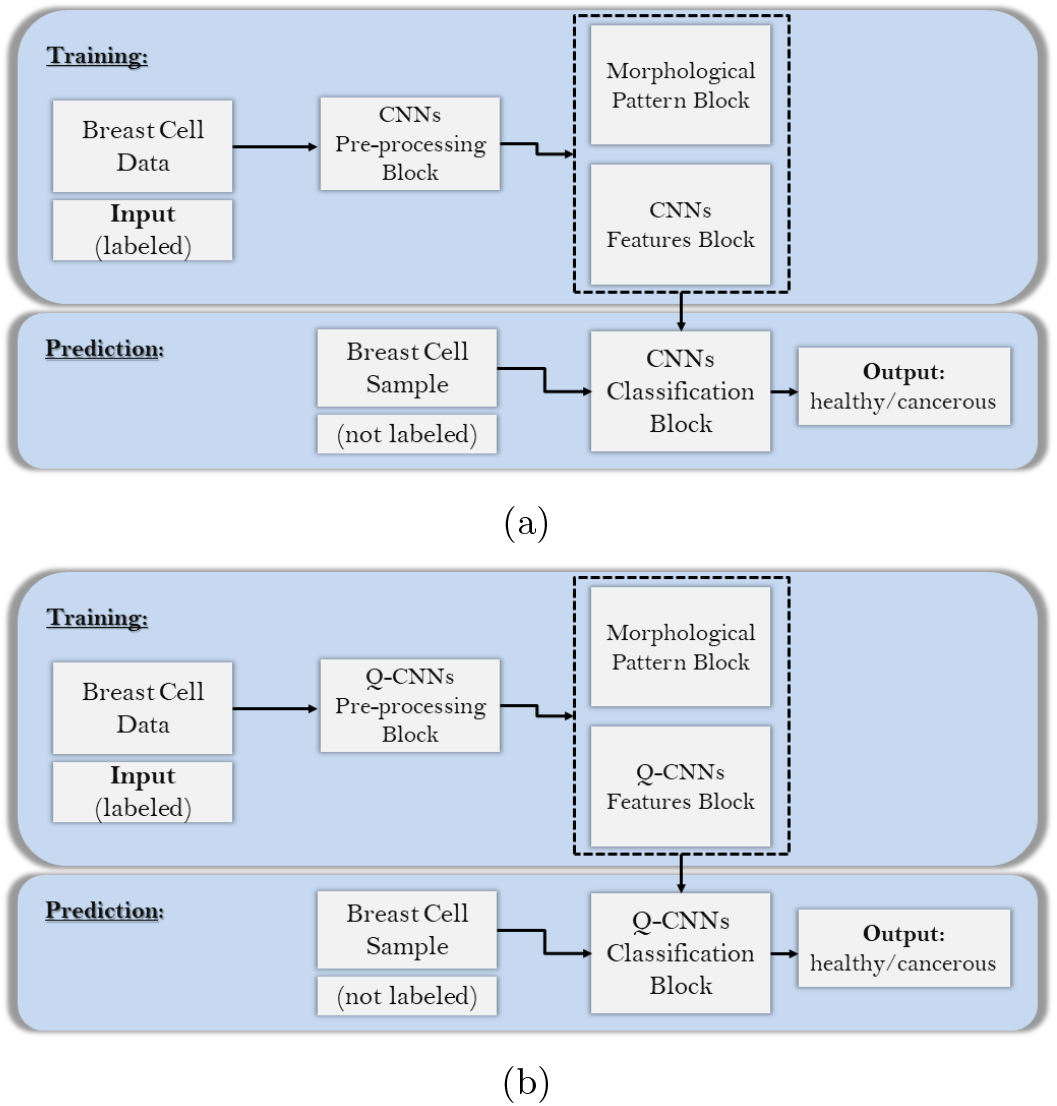
(a) Classical CNN based approach for breast cell sample. (b) Quantum CNN based approach for breast cell sample.

**FIG. 2:**
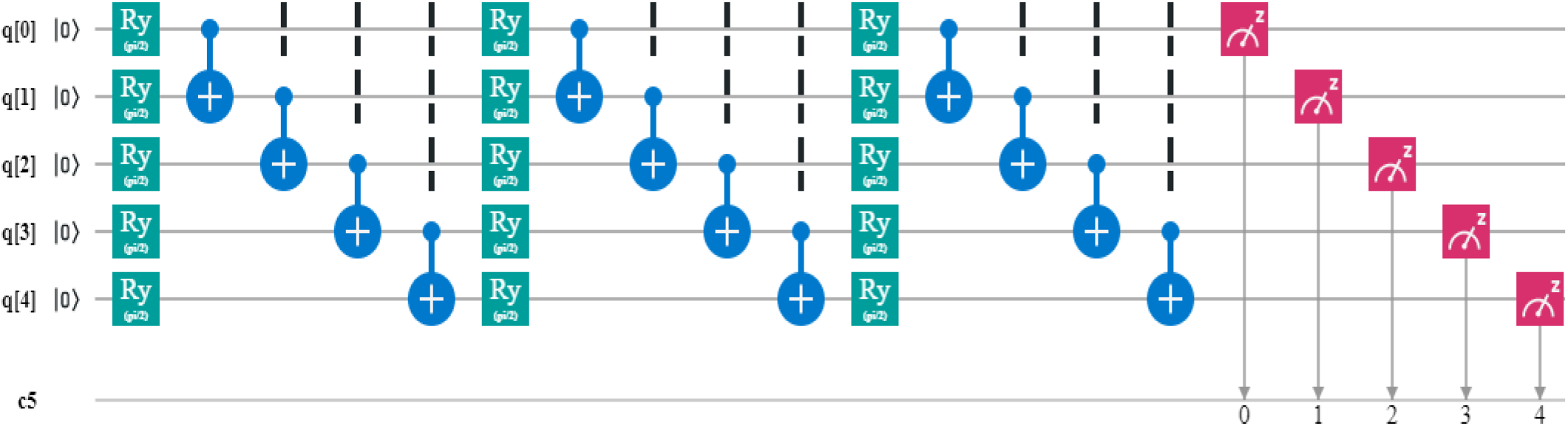
IBMQ system demonstrating how partial entanglement can be used to reduce the number of parameters required in a quantum convolutional neural network to O(*log*(*N*)).

**FIG. 3:**
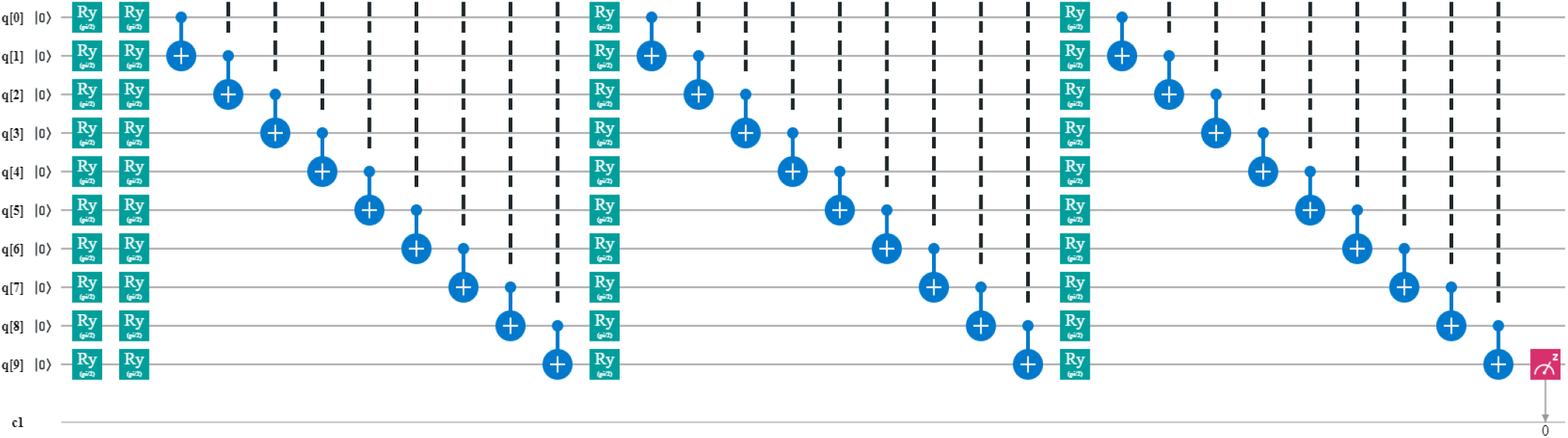
10 qubits partially entangled quantum convolutional neural network.

**FIG. 4:**
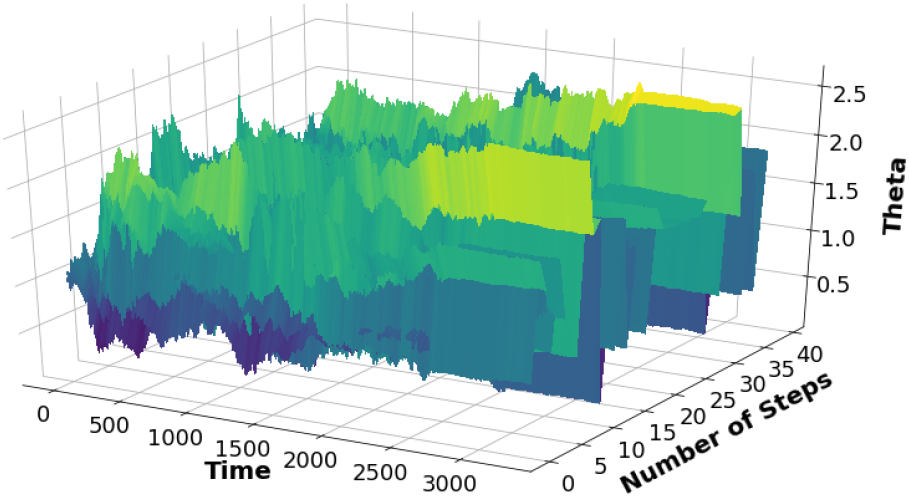
Variation in all parameters on the vertical axis. The x axis denotes the number of iterations and the y axis depicts the parameter index running from 0 to 40.

- Both CNN and QCNN input data, the first take for example an image to perform computation on it, while the second is a quantum circuit which takes in an unknown quantum state |*ψ⟩*.
- CNN relies on convolutional layers of image processing, called feature maps, each contains a number of pattern detectors *−* filters, that calculate new pixel values from the previous map by a linear combination of nearby pixels

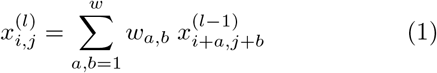

where the elements *w*_*a,b*_ from a *w×w* matrix, equivalently quantum CNN apply a layer of quasilocal unitaries on the input.

- Pooling layers, a form of operation usually added to CNN, which decreases the dimensionality of the feature maps by reducing the pixel number, QCNN does an identical work by measuring some specified qubits which collapse its states and apply rotations unitaries on the remaining ones.
- Finally, after applying a fully connected qubit unitary layer on the yielded qubits states, a measurement is acting on one qubit which output probability distribution.

Comparably to CNN, training on a large dataset optimizes the *O*(log(*N*)) parameters in the unitaries *−* N is the number of qubits, where the mean square error is calculated as:

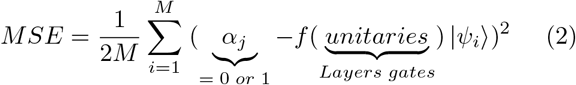

*f* (*unitaries*) |*ψ*_*i*_⟩: Denotes the expected QCNN output value for input state |*ψ*_*i*_⟩.

M: set of classified training vectors states |*ψ*_*i*_⟩, *i* = 1, …, *M* Whereas the number of those gates are fixed, they are called hyperparameters.

Interestingly, QCNN uses the same circuit structure of an important variational ansatz for many-body wavefunction *−* multiscale entanglement renormalization ansatz *−* but operate in the opposite direction. The quantum CNN has a promising application in problems of physics such as quantum phase recognition and quantum error correction optimization [20].

A CNN with *N*_*i*_ neurons in layer I and m layers have parameters, one for each connection. This goes up as O(*N* ^2^). This is where a QCNN provides an advantage. If we were to fully incorporate the effect of each qubit on the others, we would still have an O(*N* ^2^) model. This is illustrated for 5 qubits in Fig. 5. The input is 5 dimensional, each input affecting the Y rotation parameter, and the output is a 5-bit number.

**FIG. 5:**
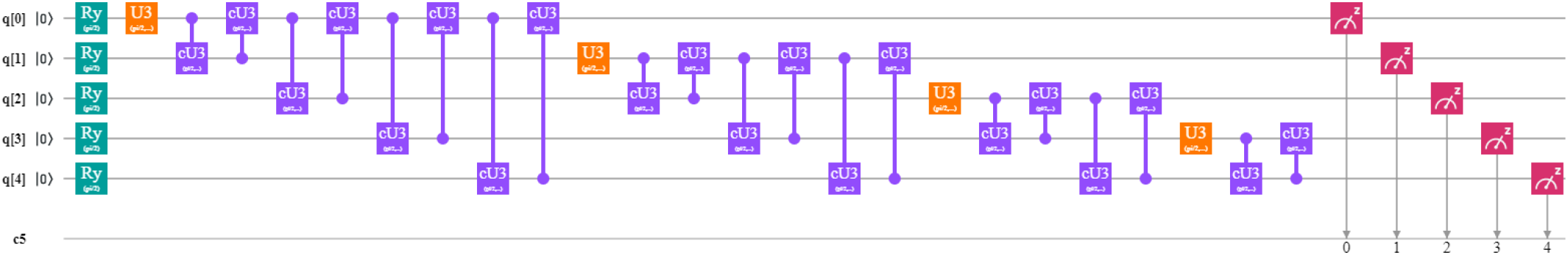
The maximally entangled qubit network is represented on IBMQ. This system scales up as O(*N* ^2^) with the number of qubits.).

Each U3 has 3 parameters, which totals to a 72-dimensional parameter space. A CNN with biases have 60 dimension parameter space. In a CNN, dropping the number of parameters results in a lack of connections between neurons, however, we can implement partial connections by entanglement in a QCNN using CX gates. Non parameterized gates allow us to entangle the inputs with lower parameter spaces. One such architecture is shown in Fig. 2. A two layer architecture only requires 10 parameters and the number of parameters go up as *O*(log(*N*)).

## IV. BREAST CANCER DETECTION

We have attempted to use our simplified QCNN to detect breast cancer as per given patient data. However, 256 qubit systems cannot be simulated efficiently on current available systems, and the exercise reduces to one of the theoretical interest for QCNNs. The breast cancer dataset of Wisconsin consists of features of lumps of the patient’s breast, e.g., radius, roughness, concavity, etc. We take 10 parameters and assign each to one qubit. The initial Y rotations are set up so that | 0⟩ corresponds to the minimum value of the variable and | 1⟩ to the maximum. As we only want to determine whether a lump is cancerous or not, we need one output bit so we only measure the last qubit’s value. We assign 0 to benign and 1 to malignant stages. The loss function is logarithmic. If *l* is the obtained label, and 1 is the expected label,

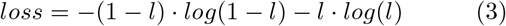

We then compute by varying each parameter one at a time. The parameters are then moved in the direction opposite the gradient, controlled by a learning rate r. As the differences in values become smaller, r is also set to smaller values.

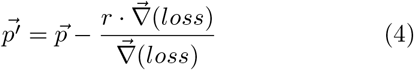

The data set is of size 600. We first take the whole data set as our training set, and then sample a subset for training, to treat the remaining as test data.

## V. RESULTS AND DISCUSSION

The simulation is carried out in two ways. The first is a test to fit the parameters to the entire data set. The risk associated to this is the over-fitting to the input set. The loss function over time is shown in Fig.7, and the final loss function over the entire data space is also depicted here. The second method involved training on a subset of the data, consisting of 100 images. After the training, the algorithm is subjected to the entire data set. This method is a more rigorous implementation of deep learning methods. The results are shown in Fig.8, and are extremely accurate considering the size of the training data set.

**FIG. 6:**
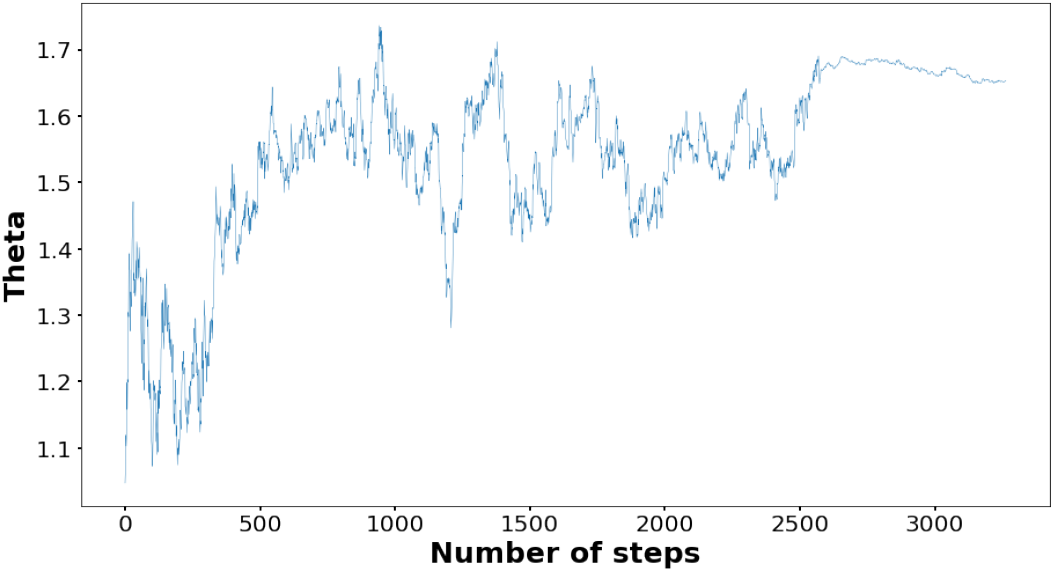
Representative variations of a parameter *θ* depicted on the y axis vs number of iterations of the simulations on the x axis. Easily visible is the definite upward trend towards a limiting value, and the point where the learning rate is manually decreased.

**FIG. 7:**
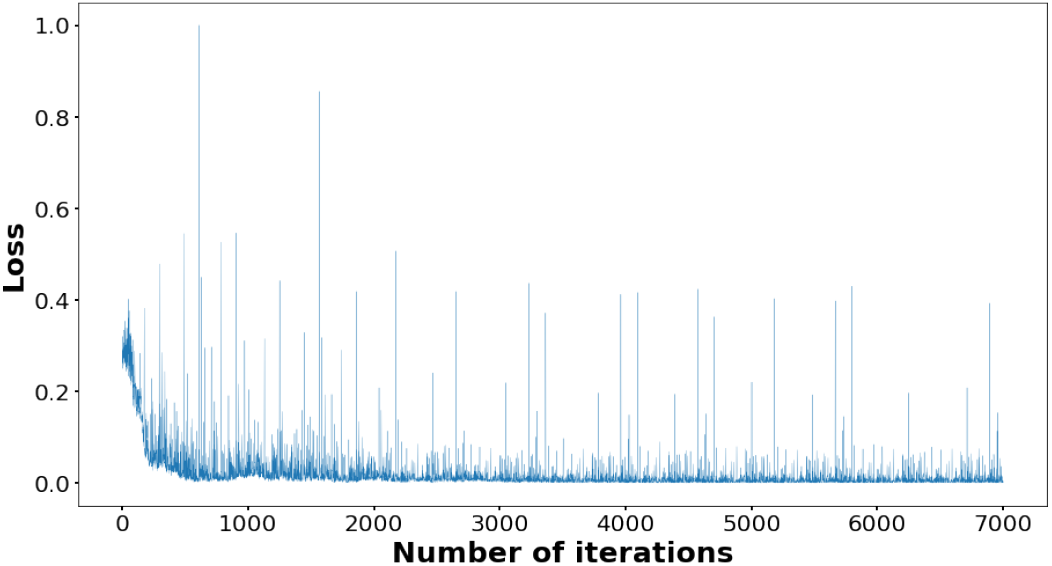
The value of the loss function depicted on the y axis vs the number of simulation iterations on the x axis. The peaks correspond to anomalous data, but the overall decrease is clearly visible.

**FIG. 8:**
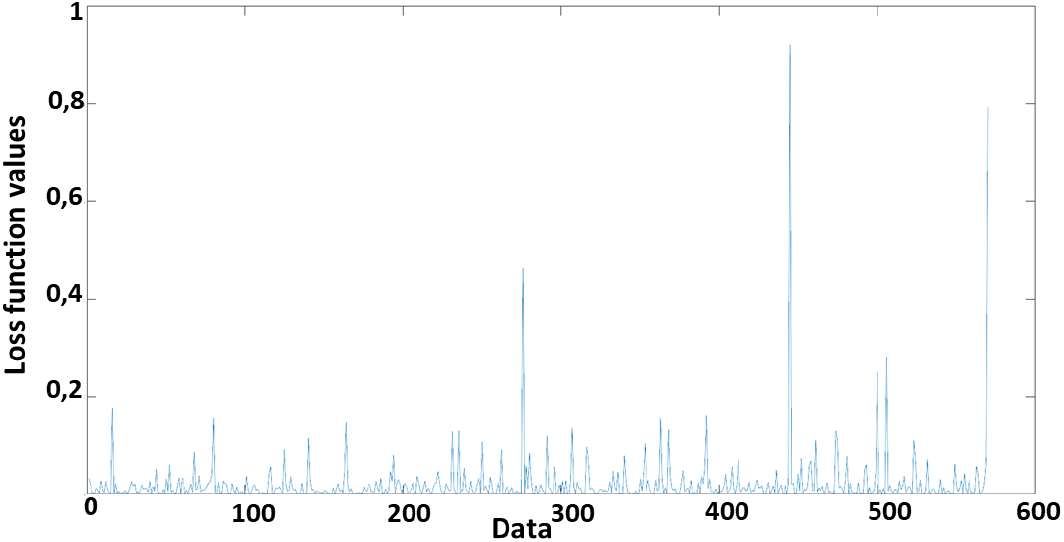
The final result for both training and testing on the entire set. The y axis is the value of the loss function for the parameter set obtained, and the x axis denotes the particular data point in the set.

The variation of the parameters over time is shown in Fig.4 and also depicted one of them as a representative parameter in Fig.6. This the point where the learning rate is manually changed and also can be observed. This is done as an emerging cyclic nature, and a change to the learning rate would disrupt the loop and allow convergence to a higher precision. The loss over most inputs slowly decreased. Some anomalous inputs resulted in wrong answers and excessive losses. The final losses for all 600 inputs are shown in Fig.9. The average loss is 1.4%. To a remarkable precision, the network is sufficiently trained in breast cancer detection.

**FIG. 9:**
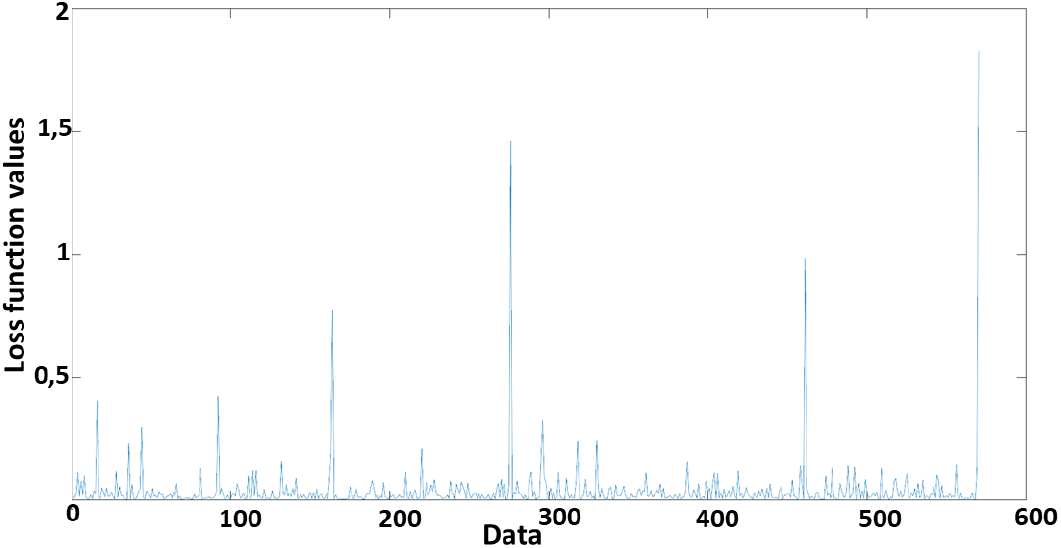
The final result when training is done on the first 100 elements and testing on the entire set. The y axis is the value of the loss function for the parameter set obtained, and the x axis denotes the particular data point in the set.

As we can see in Fig.7, the plot describes the training accuracy versus the number of training steps. Here we remark how the optimization work as expected, and like the classical case, adjusting weight and biases via back-propagation. Relying upon QCNN, this process is carried out with optimizing gates parameters using VQE algorithms where it shows a high performance regarding time complexity and accuracy.

Looking at the Fig.6, we can see that the variations of one parameter from 40 ones in the quantum circuit, where the overall variance is seen in Fig.4. We can mention about one parameter here to explain the general change. A parameter adapting to the optimum value results in the loss function convergence to the ideal value 0 as shown in Fig.7. This is achieved by an angle modification of the qubit, which attempts to shoot the correct value to eventually become stabilized, this process is worked out as follows.

We have a parameter, which is the theta in the plot. We feed an input which gives us a guess, depending on how accurate that guess is. We can change the parameters in the direction of the maximum (or minimum) gradient and shift them by a small amount, initially, the theta varies with a step size relatively big from a starting angle, this is visible in the interval between 0 and nearly 2500 on the time axis, where it fluctuates with a rapid rate, after that, we reduce this step (in the gradient function) so it takes less movement and does not over-shoot the desired value and finally, it gets balanced. Back to Fig.4, this process is done for all the remaining parameters.

Sensitivity, specificity, and accuracy are calculated using the True positive (TP), true negative (TN), false negative (FN), and false positive (FP) for making proper predictions of benign (the negative class) and malignant (the positive class) breast cell samples. From table 1, it is clearly visible that can achieve an accuracy of 98.6%, a sensitivity of 97.5%, and a specificity of 99.4% for classical CNN. Meanwhile we can observe that the QCNN can achieve an accuracy of 98.9%, a sensitivity of 97.7% and a specificity of 99.6%. Hence QCNN provides slightly better performances than the classical CNN.

**TABLE 1:** Comparative study on the performances of CNN and QCNN

|  | Accuracy | Sensitivity | Specificity |
| --- | --- | --- | --- |
| CNN | 98.6% | 97.5% | 99.4% |
| QCNN | 98.9% | 97.7% | 99.6% |

From the time complexity plot in Fig 10, it is clearly visible that the QCNN works exponentially faster than the classical algorithm.

**FIG. 10:**
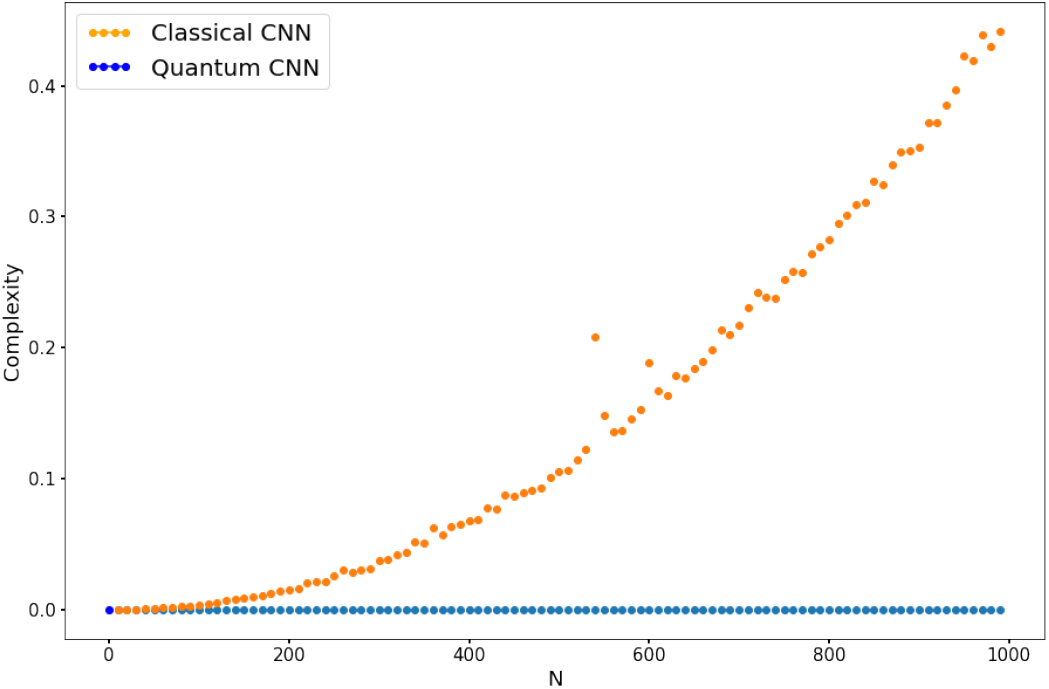
Complexity comparison between classical and quantum CNN where N denotes the number of training samples

## VI. CONCLUSION

To conclude, we have demonstrated the efficacy of quantum convolutional neural networks (QCNNs) to detect breast cancer cells. Using the proposed algorithms, we have run the circuit for different parameters and optimized it for the accurate predictions of the breast cancer cell. Using the techniques of deep learning and supervised learning in the quantum framework, we have proposed and tested the effectiveness of quantum CNN on the quantum simulator available at the IBM quantum experience platform. We have considered the application of breast cancer detection to explicate the working of our quantum convolutional neural network. We have trained the network of ten qubits in such a way that it can learn from the labeling of the given data set and can optimize the circuit parameters to obtain results with the minimum errors. Finally we also have pointed out that QCNN is not only optimum in terms of the accuracy in comparisons with classical CNN but produces results with better time complexity than the classical counterpart.

## Data Availability

no outside data is referred.

## VII. ACKNOWLEDGEMENTS

N.M., A.B., and S.K. acknowledge Indian Institute of Science Education and Research Kolkata for providing hospitality during the course of the project work. B.K.B. acknowledges the prestigious Prime Minister’s Research Fellowship awarded by DST, Govt. of India. A.B. acknowledges the IAS-INSA-NASI SRF program. We also want to thank Sudev Pradhan for editting our tex file. The authors acknowledge the support of IBM Quantum Experience for producing the experimental results. The views expressed are those of the authors and do not reflect the official policy or position of IBM or the IBM Quantum Experience team.

## References

[1] Y. Cao, G. G. Guerreschi, and A. Aspuru-Guzik, “Quantum Neuron: an elementary building block for machine learning on quantum computers,” arXiv:1711.11240, 2017.

[2] F. Tacchino, C. Macchiavello, D. Gerace, and D. Bajoni, “An artificial neuron implemented on an actual quantum processor,” npj Quantum Inf., vol.5, 2019.

[3] A. Narayanan and T. Menneer, “Quantum artificial neural network architectures and components,” Inf. Sci. vol.128, pp.231–255, 2000.

[4] Y. LeCun, Y. Bengio, and G. Hinton, “An artificial neuron implemented on an actual quantum processor,” Nature, vol.521, pp.436–444, 2015.

[5] M. Jain and S. K. Chaturvedi, “Quantum Computing Based Technique for Cancer Disease Detection System,” J. Comput. Sci. Syst. Biol., vol.7, pp.095–102, 2014.

[6] S. Mukhopadhyay, N. K. Das, I. Kurmi, A. Pradhan, N. Ghosh, and P.K. Panigrahi, “Tissue multifractality and hidden Markov model based integrated framework for optimum precancer detection,” J. Biomed. Opt., vol.22, no.10, pp.105005, 2017.

[7] Manabputra, B. K. Behera, and P. K. Panigrahi, “A Simulational Model for Witnessing Quantum Effects of Gravity Using IBM Quantum Computer,” Quantum Inf. Process., vol.19, no.119, 2020.

[8] N. Klco, E. F. Dumitrescu, A. J. McCaskey, T. D. Morris, R. C. Pooser, M. Sanz, E. Solano, P. Lougovski, and M. J. Savage, “Quantum-Classical Computations of Schwinger Model Dynamics using Quantum Computers,” Phys. Rev. A, vol.98, pp.032331, 2018.

[9] A. A. Zhukov, S. V. Remizov, W. V. Pogosov, and Y. E. Lozovik, “Algorithmic simulation of far-from-equilibrium dynamics using quantum computer,” Quantum Inf. Process., vol.17, 2018.

[10] S. Gangopadhyay Manabputra, B. K. Behera, and P. K. Panigrahi, “Generalization and demonstration of an entanglement-based Deutsch-Jozsa-like algorithm using a 5-qubit quantum computer,” Quantum Inf. Process., vol.17, 2017.

[11] A. R. Kalra, N. Gupta, B. K. Behera, S. Prakash, and P. K. Panigrahi, “Experimental Demonstration of the No Hiding Theorem Using a 5 Qubit Quantum Computer,” Quantum Inf. Process., vol.18, 2019.

[12] M. Swain, A. Rai, B. K. Behera, and P. K. Panigrahi, “Experimental demonstration of the violations of Mermin’s and Svetlichny’s inequalities for W and GHZ states,” Quantum Inf. Process., vol.18, 2019.

[13] E.P. DeBenedictis, “A Future with Quantum Machine Learning,” Computer, vol.51, pp.68–71, 2018.

[14] N. Mishra, M. Kapil, H. Rakesh, A. Anand, N. Mishra, A. Warke, S. Sarkar, S. Dutta, S. Gupta, A. P. Dash, R. Gharat, Y. Chatterjee, S. Roy, S. Raj, V. K. Jain, S. Bagaria, S. Chaudhary, V. Singh, R. Maji, P. Dalei, B. K. Behera, S. Mukhopadhyay, and P. K. Panigrahi, “Quantum Machine Learning: A Review and Current Status,” ICDMAI 2020: Springer Conference Proceeding,2020, DOI: 10.13140/RG.2.2.22824.72964.

[15] R. Harper and S. Flammia, “Fault tolerance in the IBM Q Experience,” Phys. Rev. Lett. vol.122, pp.080504, 2019.

[16] B. K. Behera, A. Banerjee, and P. K. Panigrahi, “Experimental realization of quantum cheque using a five-qubit quantum computer,” Quantum Inf. Process., vol.16, 2017.

[17] B. K. Behera, S. Seth, A. Das, and P. K. Panigrahi, “Experimental Demonstration of Quantum Repeater in IBM Quantum Computer,” Quantum Inf. Process., vol.18, 2019.

[18] B. K. Behera, T. Reza, A. Gupta, and P. K. Panigrahi, “Designing Quantum Router in IBM Quantum Computer,” Quantum Inf. Process., vol.18, 2019.

[19] Breast-cancer Data, https://www.kaggle.com/uciml/breast-cancer-wisconsin-data.

[20] I. Cong, S. Choi,and M.D. Lukin, “Quantum convolutional neural networks.” Nat. Phys., vol.15, pp.1273–1278, 2019.

